# Estimating the effect of lipids on IGF axis and subsequent breast cancer risk

**DOI:** 10.1101/2020.06.04.20122630

**Authors:** Vanessa Y Tan, Caroline J Bull, Kalina M Biernacka, Alexander Teumer, Laura Corbin, Tom Dudding, Eleanor Sanderson, Qibin Qi, Robert C Kaplan, Jerome I Rotter, Nele Friedrich, Uwe Völker, Julia Mayerle, Claire M Perks, Jeff MP Holly, Nicholas J Timpson

**Author notes:** **Corresponding author:** Professor Nicholas J. Timpson, Address: MRC Integrative Epidemiology Unit, University of Bristol, Oakfield House, Oakfield Grove, Bristol, BS8 2BN. Joint first (Vanessa Y Tan, Caroline J Bull).

## Abstract

Circulating lipids have been associated with breast cancer (BCa). This association may, in part, be due to an effect of lipids on insulin-like growth factors (IGFs), which have been reliably associated with BCa. In two-sample Mendelian randomization (MR) analyses, we found that low density lipoprotein (LDL-C) was associated with IGFBP-3 (beta:0.08 SD; 95%CI:0.02,0.15; *p* = 0.01, per SD increase in LDL-C) and IGFBP-3 was associated with postmenopausal BCa (OR:1.09; 95%CI:1.00,1.19; *p* = 0.05, per SD increase in IGFBP-3). We also found that triglycerides were associated with IGF-I (beta:-0.13SD; 95%CI:-0.22,-0.03, per SD increase in triglycerides) and that IGF-I was associated with overall BCa (OR:1.10;95%CI:1.02,1.18, per SD increase in IGF-I). Taken together, these results suggest that IGFBP-3 may be a potential causal step between LDL-C and postmenopausal BCa and IGF-I a potential causal for triglycerides. Our two-step MR results build on evidence linking circulating lipids and IGFs with BCa, however, multivariable MR analyses are currently unable to support this relationship due to weak instruments.

## Introduction

Breast cancer (BCa) is one of the leading causes of cancer-related deaths worldwide^1,2^. Circulating lipids including low density lipoprotein (LDL-C), high density lipoprotein (HDL-C) and triglycerides have been associated with known risk factors for BCa such as obesity^3,4^. However, evidence from observational studies investigating the relationship between lipid profile and BCa risk has been conflicting^5,6^. Statins competitively inhibit 3-hydroxy-3-methylglutaryl-coenzyme A (*HMGCR*) to reduce circulating LDL-C and are commonly prescribed for the prevention and treatment of cardiovascular disease, but have also been hypothesised to have anti-cancer effects^7^. Most observational studies have found no evidence of an association between statin use and BCa risk^8^. However, reverse causation, confounding and other biases may limit the interpretation of these observational findings. The associations, estimated from these observational studies, between statin therapy and breast cancer are particularly problematic, due to selection and so called immortal time biases^9^.

Mendelian randomization (MR) is an analytical approach where genetic variants reliably associated with risk factors of interest are used as “proxy” measures to investigate whether the effect of an exposure on an outcome is likely to be causal^10^. A recent MR study reported a positive association between LDL-C and HDL-C with overall or estrogen receptor (ER)-positive BCa risk^11^. It is becoming increasingly feasible to study drug effects in a MR framework using genetic variants located in the genes of the drug’s protein targets^12^. Genetic variants in *HMGCR*, associated with increased LDL-C levels, have been found to associate with increased overall and ER-positive BCa risk^13,14^. The mechanism by which statins protect against BCa and whether the effect differs by menopausal status remains to be elucidated.

Several randomised control trials have suggested that statin use is associated with alterations in circulating levels of insulin-like growth factors (IGFs), suggesting that perturbation of circulating lipids can alter levels of IGF axis traits^15–17^. The IGF axis includes IGF-I, IGF-II and six IGF binding proteins (IGFBPs), which are important for cancer initiation and progression through the regulation of cell growth, metabolism and survival^18–21^. A recent meta-analysis of individual data from observational studies found evidence that IGF-I was associated with increased BCa risk in both pre and postmenopausal women while IGFBP-3 was only associated with increased BCa risk in postmenopausal women^22^. However, it is unclear if a causal relationship between circulating IGFs and BCa exists, and whether estrogen receptor or menopausal status is important.

The role of the IGF axis in metabolic regulation is well-established^23–25^. Intervention studies in patients with growth hormone disorders support a link between IGF and lipid biology^26–28^. Population-based studies examining the relationship between circulating IGFs and lipid profile have yielded conflicting results^29–31^, however, these studies may be limited by their cross-sectional design. Hence, the direction of association and whether causation exists between circulating lipids and IGFs is still unclear. Given previous evidence implicating lipids and IGFs as potential modifiable risk factors for cancer^32–34^, there is motivation to assess the relationship between IGFs and lipids to investigate the mechanisms underlying BCa risk.

We aimed to investigate circulating IGFs as potential intermediates between lipid profile and BCa risk. We set out to examine the association between circulating lipid traits (LDL-C, HDL-C and TG), IGFs (IGF-I and IGFBP-3) and BCa (stratified by menopausal and ER status) using a two-step MR study design^35^.

## Materials and Methods

### Study design and data sources

To estimate the causal relationship between lipids and BCa using MR **Figure 1**,(i)), we obtained summary statistics from: i) a lipid GWAS conducted by the Global Lipids Genetic Consortium (GLGC, n = 188,577)^36^; ii) a BCa risk GWAS (overall (n = 228,951) and stratified by estrogen receptor status (n = 69,501 for ER positive BCa and n = 21,468 for ER negative BCa) conducted by the Breast Cancer Association Consortium (BCAC,^37^; and iii) a BCa risk GWAS (overall (n = 230,954) and stratified by menopausal status (premenopausal BCa, n = 59,124 for premenopausal BCa and n = 112,245 for postmenopausal BCa) conducted in UK Biobank (see **Supplementary methods**).

**Figure 1.**
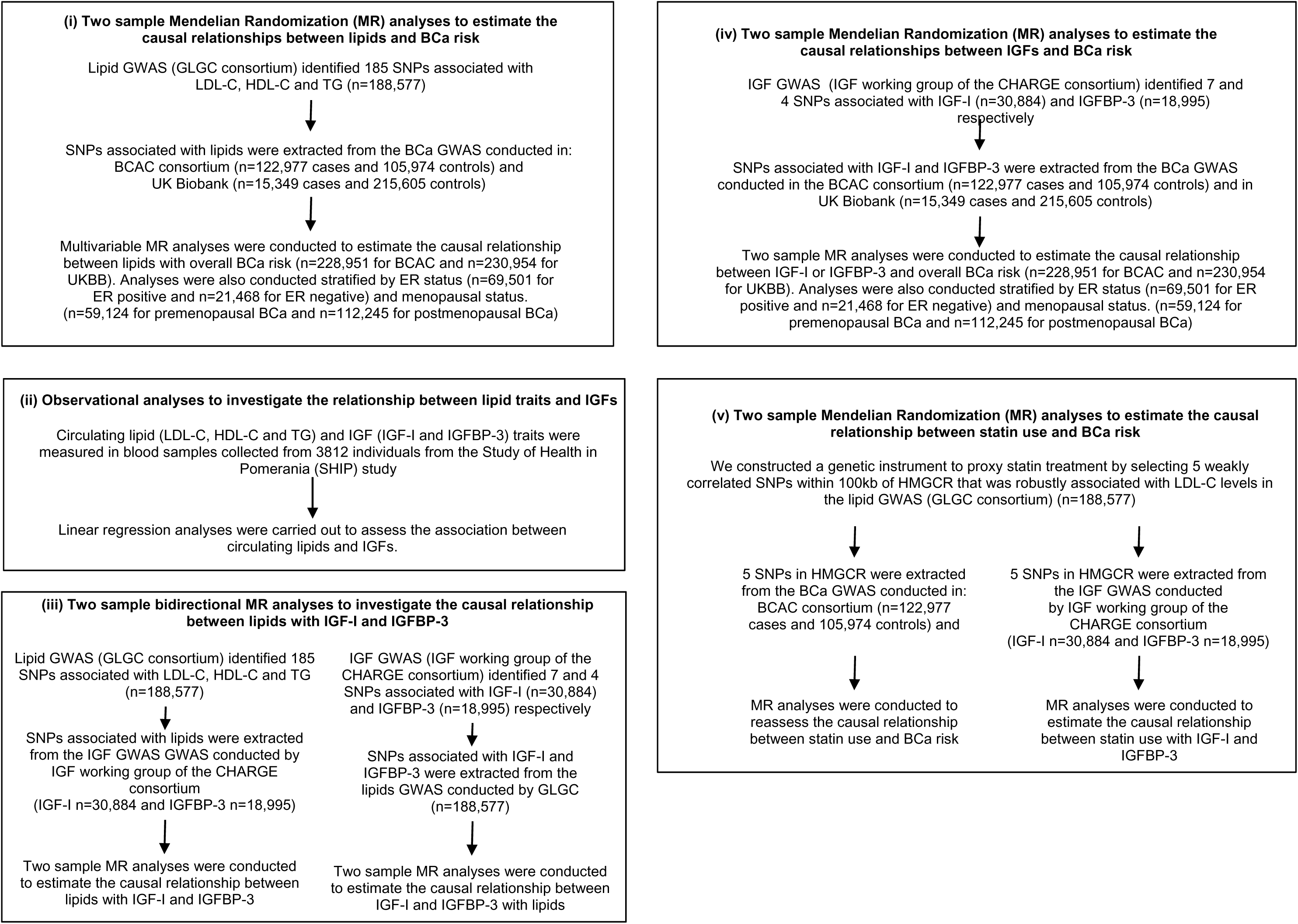
Flow diagram of study design.

To investigate the observational relationship between lipids and IGFs (**Figure 1**, (ii)), we used individual-level data (n = 3,812) from the Study of Health in Pomerania (SHIP) where circulating lipids and IGFs were measured (these data are described in detail in the **Supplementary methods**).

To estimate the causal relationship between lipids and IGF-I or IGFBP-3 using MR (**Figure 1**, (iii)), we obtained summary statistics from: i) the lipid GWAS conducted by the GLGC^36^, as described above and ii) the IGF GWAS conducted by the “IGF working group” of the CHARGE consortium (n = 30,884)^38^ (see **Supplementary methods**).

To estimate the causal relationship between circulating IGFs and BCa (**Figure 1**,(iv)), we obtained summary statistics from: i) the IGF GWAS(29); ii) the BCa risk GWAS conducted by BCAC^37^ (overall and stratified by ER status); and iii) the BCa risk GWAS conducted in UKBB (overall and stratified by menopausal status)(see **Supplementary methods**).

To assess the potential causal effect of changes in LDL-C levels due to lipid lowering drugs such as statins on BCa and IGF levels (**Figure 1**,(v)), we constructed a genetic instrument to proxy statin treatment which comprised of 5 SNPs (rs12916, rs10515198, rs12173076, rs3857388, and rs7711235) within 100kb from the gene region for *HMGCR* and in low linkage disequilibrium (r^2^< 0.20)^39^. These SNPs were robustly associated (p< 5×10^−8^) with LDL-C in the lipid GWAS conducted by GLGC^36^. We obtained summary statistics for the genetic variants in *HMGCR* from: i) the lipid GWAS [29]; ii) the IGF GWAS(29); iii) the BCa risk GWAS conducted by BCAC^37^; and iv) the BCa risk GWAS conducted in UKBB (see **Supplementary methods**).

### Genetic instruments for MR analyses

The GLGC identified 185 SNPs (r^2^ < 0.2) associated with at least HDL-C, LDL-C or TG (p< 5×10^−8^)^36^ (see **Supplementary Table 1** for list of SNPs). Due to the complex overlapping nature of the lipid traits, genetic variants are commonly associated with more than one lipid trait. To go some way to disentangling the roles of LDL-C, HDL-C and TG, we used multivariable MR which was developed to estimate the causal effect of various correlated risk factors when conditioned on one another in a single model^40,41^ (see **Supplementary methods** for more details). For the multivariable MR analyses (**Figure 1**, (i)), all 185 SNPs were included in the model. Univariable MR methods included SNPs associated with a given lipid trait at a genome-wide level of significance (p< 5×10^−8^, 76 for LDL-C, 86 for HDL-C or 51 for TG). Sex-specific GWAS results were not available for lipid traits.

For the IGF MR analyses (**Figure 1**, (iii)&(iv)), we used 7 and 4 independent SNPs (r^2^< 0.01), associated (p< 5×10^−8^) with IGF-I and IGFBP-3, respectively, as identified by a IGFs GWAS (see **Supplementary Table 2** for list of SNPs)^38^. IGF-II and IGFBP-2 were not investigated as GWAS summary results are not currently available for these traits. For IGFBP-3, marked differences in effect sizes by sex were found for 2 variants (with stronger associations in women). As differences in effect size could potentially bias MR estimates^42^, analyses estimating the causal relationship between IGF-I or IGFBP-3 and BCa were conducted using female-specific effect estimates for IGF-I and IGFBP-3.

### Statistical analyses

Multivariable MR was used to estimate the effect of LDL-C, HDL-C and TG on the odds of BCa when conditioned on one another (**Figure 1**, (i)). As there were likely to be differences between the causal estimates between BCAC and UKBB, a random-effects meta-analysis was used to combine MR estimates using BCAC and UKBB summary statistics using the ‘metan’ package in Stata. Analyses stratified by ER status (in BCAC) and menopausal status (in UKBB) were also conducted.

Observational associations between lipids and IGF-I or IGFBP-3 were assessed using linear regression (**Figure 1**, (ii)). Fully adjusted models included age, sex, smoking status, body mass index (BMI) and diabetes status. Associations of lipids and IGFs with potential confounders were estimated using linear regression.

Bidirectional MR was performed to assess the causal effect of lipids on IGF-I and IGFBP-3 and vice versa (**Figure 1**, (iii)). SNP-exposure and SNP-outcome associations were combined using the inverse variance weighted (IVW) method^43^. MR-Egger^44^, weighted median^45^ and multivariable MR^40,41^ were performed as sensitivity analyses.

To assess the causal effect of IGFs on BCa, SNP-exposure (SNP-IGF) and SNP-outcome (SNP-BCa) associations were combined using inverse variance weighting (**Figure 1**, (iv))^43^. MR-Egger and weighted median were not conducted as these lack power for instruments with a small number of SNPs^46^. SNPs associated with IGF-I are also associated with other components of the IGF pathway^38^. To attempt to account for the overlapping associations for these SNPs, we employed multivariable MR by regression of the SNP-BCa estimates on SNP-IGF-I and SNP-IGFBP-3 estimates in a multivariable weighted regression model. A random-effects meta-analysis was used to combine BCAC and UKBB estimates. Analyses stratified by ER status (in BCAC) and menopausal status (in UKBB) were also conducted.

Multivariable MR^47^ was conducted as an extension of the IVW method to test the hypothesis that circulating IGF-I or IGFBP3 act as intermediate factors in any association between lipid profile and BCa, identified by the analyses outlined above (**Figure 1**, (i-iv)). Two-sample Conditional F-statistics^48^ were estimated to predict instrument strength in the multivariable model.

To estimate the causal relationship between statin use and BCa or IGF levels, we used a random-effect linear regression model weighted by the inverse variance of the SNP-BCa associations. The genetic correlation between the 5 SNPs in *HMGCR* was incorporated into the MR model using the weighted generalized linear regression method implemented in the ‘MendelianRandomization’ R package^49,50^.

All MR analyses were performed using the MR-Base “TwoSampleMR” package^51^. All other statistical analyses were performed using Stata version 14 (StataCorp, College Station, Texas, USA) or R version 3.2.4.

## Results

### Re-assessing the causal relationship between circulating lipids with breast cancer

The relationship between circulating lipids and BCa in BCAC and UKBB was estimated using multivariable MR. In BCAC, the odds ratio (OR) for overall BCa per SD increase in LDL-C, HDL-C and TG was 1.06 (95%CI 1.01, 1.12; *p* = 0.02), 1.07 (95%CI 1.01, 1.14; *p* = 0.03) and 0.93 (95%CI: 0.86, 1.00; *p* = 0.06), respectively (**Figure 2**). In UKBB, MR estimates of the association between LDL-C, HDL-C, and TG and BCa were consistent with results in BCAC (**Figure 2**). In a meta-analysis of BCAC and UKBB data, the OR for overall BCa per SD increase in LDL-C, HDL-C and TG was 1.07 (95%CI 1.02, 1.12; *p* = 0.01), 1.06 (95%CI 1.01, 1.10; *p* = 0.01) and 0.94 (95%CI 0.89, 1.00; *p* = 0.05), respectively (**Figure 2**).

**Figure 2.**
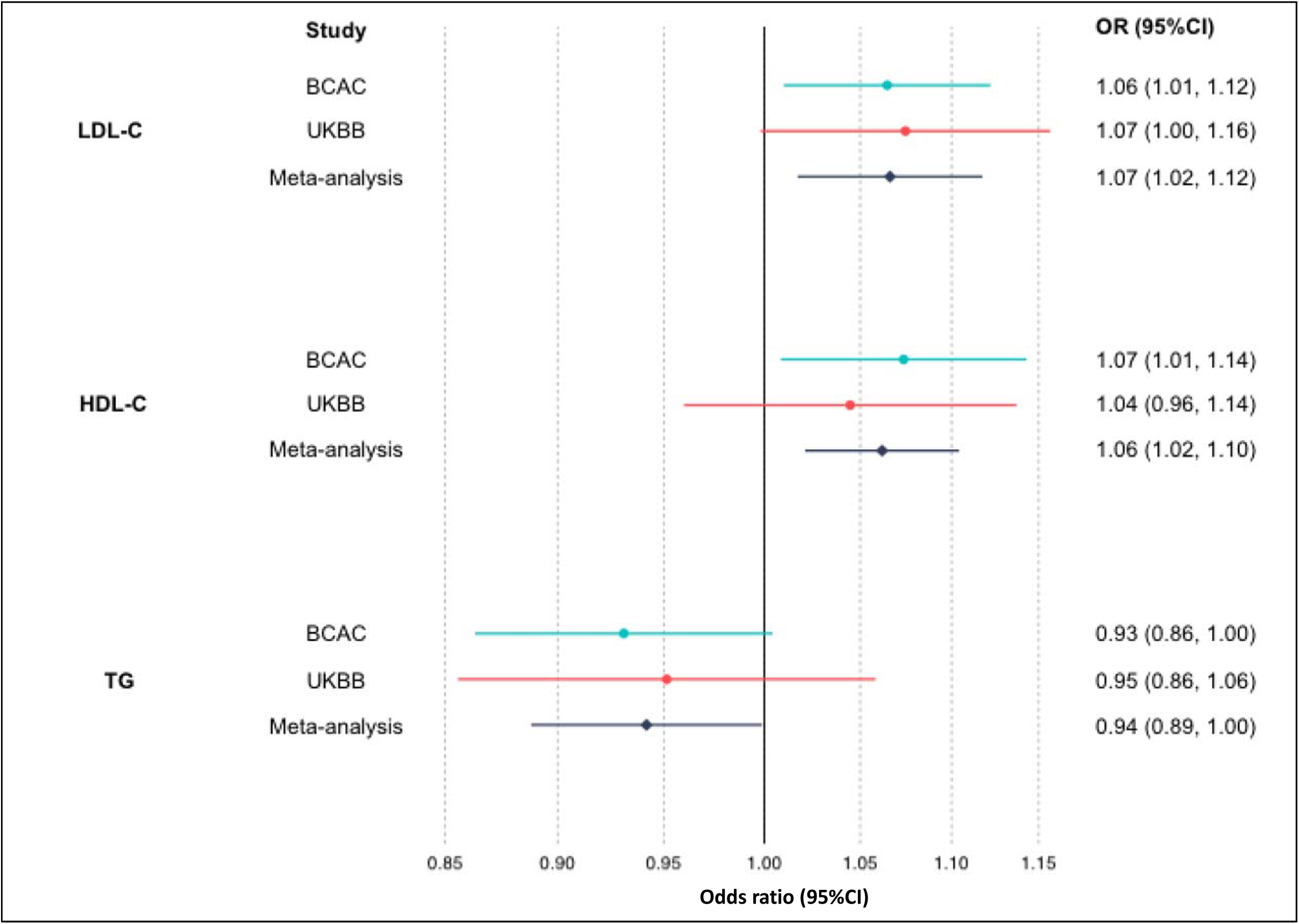
Estimates of the causal relationship between circulating lipid traits and breast cancer risk in BCAC and UK Biobank. The forest plot shows the estimate of the causal effect of LDL-C, HDL-C and TG on overall breast cancer risk using summary data from the breast cancer association consortium (BCAC) (n=122,977 cases and 105,974 controls) and UK Biobank (UKBB) (n = 15,349 cases and 215,605 controls). Squares represent point estimates from individual analyses. Horizontal lines represent the 95% confidence intervals. Pooled estimates (represented by diamonds) were obtained using random-effects meta-analysis.

Using data from BCAC, we extended this to the investigation of the relationship between circulating lipids and BCa, stratified by ER status. The OR for ER-positive BCa per SD increase in LDL-C, HDL-C and TG was 1.06 (95%CI 0.99, 1.12; *p* = 0.07), 1.08 (95%CI 1.01, 1.15; *p* = 0.04) and 0.93 (95%CI 0.86, 1.02; *p* = 0.11), respectively. The OR for ER-negative BCa per SD increase in LDL-C, HDL-C and TG was 1.05 (95%CI 0.98, 1.13; *p* = 0.16), 1.08 (95%CI 0.99, 1.18; *p* = 0.09) and 0.92 (95%CI 0.83, 1.02; *p* = 0.11), respectively (**Table 1**).

**Table 1.** Estimates of the causal relationship between lipids and breast cancer stratified by ER status or menopausal status.

| Exp | Consortium | Outcome | N | OR<sup>a</sup><br>(95% CI) | p |
| --- | --- | --- | --- | --- | --- |
| **LDL-C** | BCAC | ER positive BCa | 69,501 | 1.06<br>(0.99, 1.12) | 0.07 |
|  | BCAC | ER negative BCa | 21,468 | 1.05<br>(0.98, 1.13) | 0.16 |
| **HDL-C** | BCAC | ER positive BCa | 69,501 | 1.08<br>(1.01, 1.15) | 0.04 |
|  | BCAC | ER negative BCa | 21,468 | 1.08<br>(0.99, 1.18) | 0.09 |
| **TG** | BCAC | ER positive BCa | 69,501 | 0.93<br>(0.86, 1.02) | 0.11 |
|  | BCAC | ER negative BCa | 21,468 | 0.92<br>(0.83, 1.02) | 0.11 |
| **LDL-C** | UKBB | Premenopause BCa | 59,124 | 1.07<br>(0.91, 1.26) | 0.42 |
|  | UKBB | Postmenopause BCa | 112,245 | 1.14<br>(1.04, 1.25) | 0.004 |
| **HDL-C** | UKBB | Premenopause BCa | 59,124 | 1.07<br>(0.89, 1.30) | 0.46 |
|  | UKBB | Postmenopause BCa | 112,245 | 1.02<br>(0.93, 1.13) | 0.64 |
| **TG** | UKBB | Premenopause BCa | 59,124 | 1.02<br>(0.81, 1.30) | 0.84 |
|  | UKBB | Postmenopause BCa | 112,245 | 0.91<br>(0.80, 1.03) | 0.14 |

OR, odds ratio; 95% CI, 95% confidence interval; *p*, p-value; HDL-C, high density lipoprotein cholesterol; LDL-C, low density lipoprotein cholesterol; TG, triglycerides; MR, Mendelian randomization; BCa, Breast cancer; BCAC, breast cancer association consortium; UKBB, UK Biobank; ER, estrogen receptor.
<sup>a</sup>Associations are per 1 SD unit increase in LDL-C, HDL-C or TG from multivariable MR analyses

Using data from UKBB, we investigated the relationship between circulating lipids and BCa, stratified by menopausal status. We found evidence that LDL-C was associated with increased odds of postmenopausal BCa (OR 1.14, 95%CI 1.04, 1.25; *p* = 0.004), but there was weak evidence for premenopausal BCa (OR 1.07, 95%CI 0.91, 1.26; *p* = 0.42). There was little evidence to suggest that HDL-C or TG associated with either premenopausal or postmenopausal BCa (**Table 1**).

### Observational relationship between circulating lipids and levels of IGF-I and IGFBP-3 in the SHIP study

Study characteristics of the SHIP study are shown in **Supplementary Table 3**. The mean (SD) IGF-I and IGFBP-3 levels were 142.1 (57.6) ng/ml and 1884.9 (503.5) ng/ml, respectively. The mean (SD) LDL-C, HDL-C and TG levels were 3.57 (1.16) mmol/l, 1.45 (0.44) mmol/l and 1.82 (1.30) mmol/l, respectively. For IGF-I, a SD unit increase in LDL-C, HDL-C and TG was associated with a –0.11 (95%CI –0.14, –0.08; *p* = 2.27×10^−12^), 0.02 (95%CI –0.01, 0.05; *p* = 0.29) and –0.16 (95%CI –0.19, –0.13; *p* = 2.26×10^−22^) SD unit change in IGF-I levels, respectively. For IGFBP-3, a SD unit increase in LDL-C, HDL-C and TG was associated with a –0.01 (95%CI –0.04, 0.02; *p* = 0.47), 0.06 (95%CI 0.03, 0.10; *p* = 7.01×10^−5^) and 0.03 (95%CI –0.003, 0.06; *p* = 0.08) SD unit change in IGFBP-3 levels, respectively. Circulating lipids and IGF traits were associated with potential confounders of a lipid- or IGF-BCa relationship, including age, sex, smoking status, diabetes status and body mass index(**Supplementary Table 4**). In the fully adjusted model, a SD unit increase in LDL-C, HDL-C and TG was associated with a 0.03 (95%CI 0.004, 0.06; *p* = 0.03), –0.05 (95%CI –0.08, –0.02; *p* = 0.001) and –0.06 (95%CI –0.09, –0.04; *p* = 1.5×10^−5^) SD unit change in IGF-I levels, respectively. For IGFBP-3, a SD unit increase in LDL-C, HDL-C and TG was associated with a 0.08 (95%CI 0.05, 0.11; *p* = 1.12×10^−7^), –0.02 (95%CI –0.05, 0.02; *p* = 0.30) and 0.13 (95%CI 0.10, 0.16; *p* = 2.69×10^−15^) SD unit change in IGFBP-3 levels, respectively (**Table 2**).

**Table 2.** Beta estimates of SD unit change in IGF-I and IGFBP-3 per SD unit increase in HDL-C, LDL-C or TG based on observational analyses in SHIP.

| Exposure | Outcome | N | Unadjusted |  | Adjusted<sup>a</sup> |  |
| --- | --- | --- | --- | --- | --- | --- |
|  |  |  | Beta<sup>b</sup><br>(95%CI) | p | Beta<sup>b</sup><br>(95%CI) | p |
| LDL-C | IGF-I | 3812 | -0.11<br>(-0.14, -0.08) | 2x10<sup>-12</sup> | 0.03<br>(0.004, 0.06) | 0.03 |
| HDL-C | IGF-I | 3812 | 0.02<br>(-0.01, 0.05) | 0.29 | -0.05<br>(-0.08, -0.02) | 0.001 |
| TG | IGF-I | 3812 | -0.16<br>(-0.19, -0.13) | 2x10<sup>-22</sup> | -0.06<br>(-0.09, -0.04) | 1.5x10<sup>-5</sup> |
| LDL-C | IGFBP-3 | 3812 | -0.01<br>(-0.04, 0.02) | 0.47 | 0.08<br>(0.05, 0.11) | 1.12x10<sup>-7</sup> |
| HDL-C | IGFBP-3 | 3812 | 0.06<br>(0.03, 0.10) | 7.01x10<sup>-5</sup> | -0.02<br>(-0.05, 0.02) | 0.30 |
| TG | IGFBP-3 | 3812 | 0.03<br>(-0.003, 0.06) | 0.08 | 0.13<br>(0.10, 0.16) | 2.69x10<sup>-15</sup> |

N, sample size; Beta, regression coefficient; 95% CI, 95% confidence interval; p, p-value; HDL-C, high density lipoprotein cholesterol; LDL-C, low density lipoprotein cholesterol; TG, triglycerides; IGF-I, insulin-like growth factor 1; IGFBP-3, insulin-like growth factor binding protein 3
<sup>a</sup>Analyses were adjusted for age, gender, smoking status, body mass index and diabetes status
<sup>b</sup>Beta refers to the s.d unit change in IGF-I or IGFBP-3 levels per 1 s.d unit decrease in HDL-C, LDL-C or TG.

### Estimating the causal relationship between circulating lipids and IGFs

There was little evidence to suggest that LDL-C or HDL-C alter IGF-I levels. The MR estimate of the causal relationship between TG and IGF-I was –0.13 SD units (95%CI –0.22, –0.03; *p* = 0.01) per SD unit increase in TG. Estimates obtained using MR-Egger regression, weighted median and multivariable MR methods were of similar magnitude (**Table 3**). There was little evidence for directional pleiotropy using Egger regression (Intercept: 0.007; se: 0.004; directionality *p* = 0.11). In the reverse direction, there was little evidence to suggest that IGF-I altered circulating LDL-C, HDL-C or TG (**Table 4**).

**Table 3.** Beta estimates of SD unit change in IGF-I and IGFBP-3 per SD unit increase in HDL-C, LDL-C or TG based on two-sample and multivariable Mendelian randomization analyses.

| Exposure | Outcome | Main analysis |  | Sensitivity analyses |  |  |  |  |  |
| --- | --- | --- | --- | --- | --- | --- | --- | --- | --- |
|  |  | IVW<br>Beta<sup>a</sup><br>(95% CI) | *p* | Weighted<br>Median<br>Beta<sup>a</sup><br>(95% CI) | *p* | MR Egger<br>regression<br>Beta<sup>a</sup><br>(95% CI) | *p* | Multivariable<br>MR<br>Beta<sup>a</sup><br>(95% CI) | *p* |
| LDL-C | IGF-I | 0.01<br>(-0.04, 0.07) | 0.64 | 0.02<br>(-0.05, 0.09) | 0.64 | 0.001<br>(-0.09, 0.10) | 0.99 | 0.04<br>(-0.02, 0.10) | 0.21 |
| HDL-C | IGF-I | 0.01<br>(-0.05, 0.07) | 0.80 | -0.01<br>(-0.08, 0.07) | 0.87 | 0.03<br>(-0.08, 0.13) | 0.62 | -0.02<br>(-0.08, 0.04) | 0.56 |
| TG | IGF-I | -0.13<br>(-0.22, -0.03) | 0.01 | -0.04<br>(-0.13, 0.06) | 0.46 | -0.23<br>(-0.39, -0.07) | 0.01 | -0.12<br>(-0.18, -0.06) | <0.001 |
| LDL-C | IGFBP-3 | 0.08<br>(0.02, 0.15) | 0.01 | 0.05<br>(-0.03, 0.12) | 0.26 | 0.09<br>(-0.02, 0.20) | 0.10 | 0.08<br>(0.02, 0.14) | 0.01 |
| HDL-C | IGFBP-3 | -0.06<br>(-0.13, 0.01) | 0.08 | -0.05<br>(-0.17, 0.06) | 0.36 | -0.05<br>(-0.17, 0.06) | 0.36 | -0.02<br>(-0.08, 0.04) | 0.61 |
| TG | IGFBP-3 | 0.10<br>(0.01, 0.19) | 0.03 | 0.08<br>(-0.03, 0.19) | 0.14 | 0.13<br>(-0.02, 0.27) | 0.10 | 0.06<br>(0.001, 0.12) | 0.04 |

N, sample size; Beta, regression coefficient; IVW, inverse variance weighted; 95% CI, 95% confidence interval; *p*, *p*-value; MR, mendelian randomization; HDL-C, high density lipoprotein cholesterol; LDL-C, low density lipoprotein cholesterol; TG, triglycerides
<sup>a</sup>Beta refers to the s.d unit change in IGF-I or IGFBP-3 levels per 1 s.d unit increase in HDL-C, LDL-C or TG.

**Table 4.** Beta estimates of SD unit change in LDL-C, HDL-C and triglycerides per SD unit increase in IGF-I or IGFBP-3 based on two-sample and multivariable Mendelian randomization analyses.

| Exposure | Outcome | Main analysis |  | Sensitivity analysis |  |
| --- | --- | --- | --- | --- | --- |
|  |  | IVW<br>Beta<sup>a</sup><br>(95% CI) | p | Multivariable MR<br>Beta<sup>a</sup><br>(95% CI) | p |
| IGF-I | LDL-C | -0.07<br>(-0.21, 0.08) | 0.37 | -0.04<br>(-0.18, 0.10) | 0.55 |
| IGF-I | HDL-C | -0.02<br>(-0.13, 0.08) | 0.65 | -0.01<br>(-0.11, 0.09) | 0.77 |
| IGF-I | TG | -0.39<br>(-1.00, 0.22) | 0.21 | -0.35<br>(-0.90, 0.20) | 0.20 |
| IGFBP-3 | LDL-C | -0.01<br>(-0.04, 0.02) | 0.60 | -0.01<br>(-0.07, 0.05) | 0.81 |
| IGFBP-3 | HDL-C | -0.002<br>(-0.02, 0.02) | 0.87 | -0.01<br>(-0.05, 0.03) | 0.69 |
| IGFBP-3 | TG | -0.02<br>(-0.05, 0.02) | 0.33 | 0.02<br>(-0.22, 0.26) | 0.90 |

N, sample size; Beta, regression coefficient; IVW, inverse variance weighted; 95% CI, 95% confidence interval; p, p-value; HDL-C, high density lipoprotein cholesterol; LDL-C, low density lipoprotein cholesterol; TG, triglycerides
<sup>a</sup>Beta refers to the s.d unit change in HDL-C, LDL-C, TG or TC per SD unit change in IGF-I and IGFBP-3 levels.

The MR estimate of the causal relationship between LDL-C and IGFBP-3 was 0.08 SD units (95%CI 0.02, 0.15; *p* = 0.01) per SD unit increase in LDL. There was little evidence for an association between HDL-C and IGFBP-3 (**Table 3**). The IVW estimate of the causal relationship between TG and IGFBP-3 was 0.10 SD units (95%CI 0.01, 0.19; *p* = 0.03) per SD unit increase in TG. Estimates obtained using MR-Egger regression, weighted median and multivariable MR methods were consistent with the IVW analyses for LDL-C and TG (**Table 3**). In the reverse direction, there was minimal evidence to suggest that IGFBP-3 altered circulating LDL-C, HDL-C or TG (**Table 4**).

### Estimating the causal relationship between circulating IGFs and breast cancer

Using data from BCAC, the OR for overall BCa per SD increase in IGF-I and IGFBP-3 (using the IVW method) was 1.10 (95%CI 1.01, 1.20; *p* = 0.02) and 1.00 (95%CI 0.96, 1.03; *p* = 0.95), respectively. Estimates from multivariable MR analyses were consistent with the IVW analyses unadjusted for IGF-I (OR 1.12, 95%CI 1.03, 1.20; *p* = 0.01) and IGFBP-3 (OR 0.98, 95%CI 0.95, 1.02; *p* = 0.40). In UKBB, estimates for the effect of IGF-I on BCa were comparable. For IGFBP-3, the OR for overall BCa per SD increase in IGFBP-3 was 1.08 (95%CI 1.01, 1.15; *p* = 0.03) and 1.08 (95%CI 1.01, 1.16; *p* = 0.02), estimated by IVW and multivariable MR, respectively (**Table 5**). When results from BCAC and UKBB were meta-analysed, the OR for overall BCa for IGF-I and IGFBP-3 was 1.10 (95%CI 1.02, 1.18; *p* = 0.01) and 1.02 (95%CI 0.99, 1.06; *p* = 0.21), respectively, from IVW analyses (**Figure 3**).

**Table 5.**
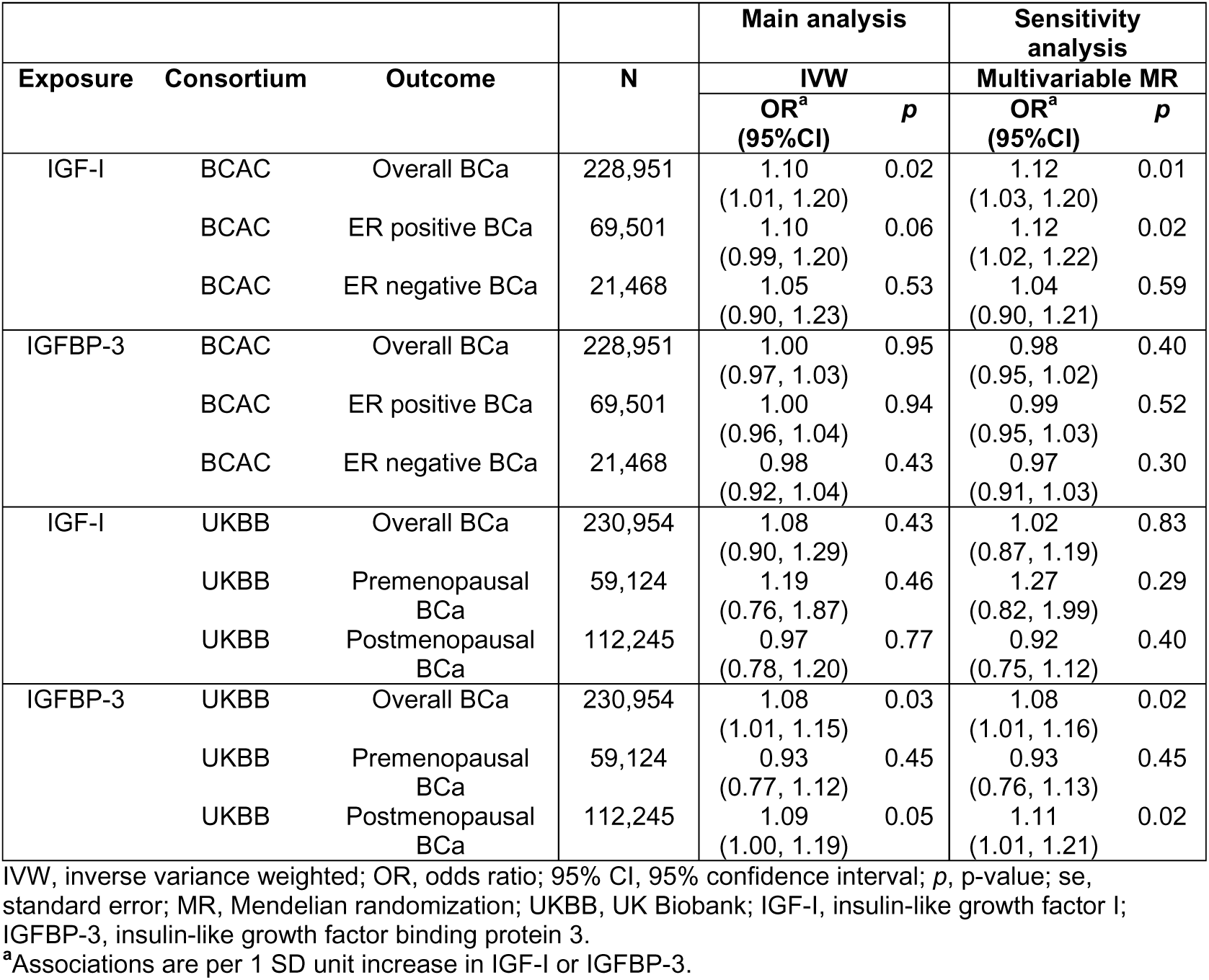
Estimates of the causal relationship between IGF-I or IGFBP-3 and breast cancer stratified by ER or menopausal status.

| Exposure | Consortium | Outcome | N | Main analysis |  | Sensitivity analysis |  |
| --- | --- | --- | --- | --- | --- | --- | --- |
|  |  |  |  | IVW |  | Multivariable MR |  |
|  |  |  |  | OR<sup>a</sup><br>(95%CI) | p | OR<sup>a</sup><br>(95%CI) | p |
| IGF-I | BCAC | Overall BCa | 228,951 | 1.10<br>(1.01, 1.20) | 0.02 | 1.12<br>(1.03, 1.20) | 0.01 |
|  | BCAC | ER positive BCa | 69,501 | 1.10<br>(0.99, 1.20) | 0.06 | 1.12<br>(1.02, 1.22) | 0.02 |
|  | BCAC | ER negative BCa | 21,468 | 1.05<br>(0.90, 1.23) | 0.53 | 1.04<br>(0.90, 1.21) | 0.59 |
| IGFBP-3 | BCAC | Overall BCa | 228,951 | 1.00<br>(0.97, 1.03) | 0.95 | 0.98<br>(0.95, 1.02) | 0.40 |
|  | BCAC | ER positive BCa | 69,501 | 1.00<br>(0.96, 1.04) | 0.94 | 0.99<br>(0.95, 1.03) | 0.52 |
|  | BCAC | ER negative BCa | 21,468 | 0.98<br>(0.92, 1.04) | 0.43 | 0.97<br>(0.91, 1.03) | 0.30 |
| IGF-I | UKBB | Overall BCa | 230,954 | 1.08<br>(0.90, 1.29) | 0.43 | 1.02<br>(0.87, 1.19) | 0.83 |
|  | UKBB | Premenopausal BCa | 59,124 | 1.19<br>(0.76, 1.87) | 0.46 | 1.27<br>(0.82, 1.99) | 0.29 |
|  | UKBB | Postmenopausal BCa | 112,245 | 0.97<br>(0.78, 1.20) | 0.77 | 0.92<br>(0.75, 1.12) | 0.40 |
| IGFBP-3 | UKBB | Overall BCa | 230,954 | 1.08<br>(1.01, 1.15) | 0.03 | 1.08<br>(1.01, 1.16) | 0.02 |
|  | UKBB | Premenopausal BCa | 59,124 | 0.93<br>(0.77, 1.12) | 0.45 | 0.93<br>(0.76, 1.13) | 0.45 |
|  | UKBB | Postmenopausal BCa | 112,245 | 1.09<br>(1.00, 1.19) | 0.05 | 1.11<br>(1.01, 1.21) | 0.02 |

IVW, inverse variance weighted; OR, odds ratio; 95% CI, 95% confidence interval; p, p-value; se, standard error; MR, Mendelian randomization; UKBB, UK Biobank; IGF-I, insulin-like growth factor I; IGFBP-3, insulin-like growth factor binding protein 3.
<sup>a</sup>Associations are per 1 SD unit increase in IGF-I or IGFBP-3.

**Figure 3.**
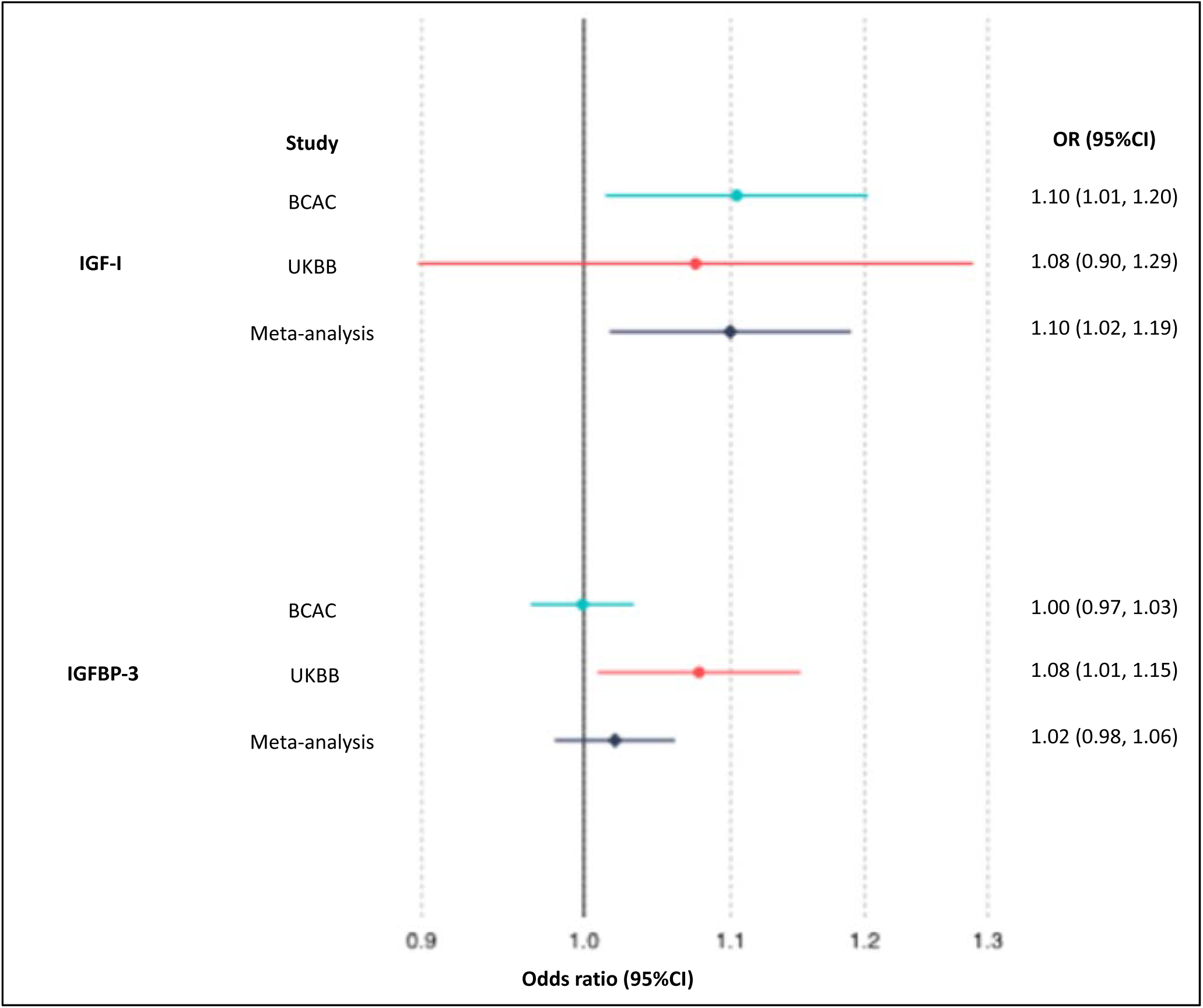
Estimates of the causal relationship between circulating IGF traits and breast cancer risk in BCAC and UK Biobank. The forest plot shows the estimate of the causal effect of IGF-I and IGFBP-3 on breast cancer risk using summary data from the breast cancer association consortium (BCAC) (n = 122,977 cases and 105,974 controls) and UK Biobank (UKBB) (n = 15,349 cases and 215,605 controls). Squares represent point estimates from individual analyses. Horizontal lines represent the 95% confidence intervals. Pooled estimates (represented by diamonds) were obtained using random effects meta-analysis.

We also investigated the relationship between IGF traits and BCa by ER-status. The OR for ER-positive BCa per SD increase in IGF-I and IGFBP-3 (using the IVW method) was 1.10 (95%CI: 0.99, 1.20; *p* = 0.06) and 1.00 (95%CI: 0.96, 1.04; *p* = 0.94), respectively. The OR for ER-negative BCa per SD increase in IGF-I and IGFBP-3 (using the IVW method) was 1.05 (95%CI: 0.90, 1.23; *p* = 0.53) and 0.98 (95%CI: 0.92, 1.04; *p* = 0.43), respectively. For IGF-I and IGFBP-3, multivariable MR and IVW estimates were consistent for both ER-positive and ER-negative BCa (**Table 5**).

Using data from UKBB, we investigated the relationship between IGF traits and BCa stratified by menopausal status. The OR for postmenopausal BCa per SD increase in IGF-I and IGFBP-3 (using the IVW method) was 0.97 (95%CI 0.78, 1.20; *p* = 0.77) and 1.09 (95%CI 1.00, 1.19; *p* = 0.05), respectively. The OR for premenopausal BCa per SD increase in IGF-I and IGFBP-3 (using the IVW method) was 1.19 (95%CI 0.76, 1.87; *p* = 0.46) and 0.93 (95%CI 0.77, 1.12; *p* = 0.45), respectively. For IGF-I and IGFBP-3, multivariable MR and IVW estimates were consistent for both pre- and post-menopausal BCa (**Table 5**).

### Multivariable MR analyses to estimate whether IGFs are on the causal pathway between lipids and BCa

As a sensitivity analysis, MVMR analyses were conducted to investigate if the effect of TG on overall BCa was attenuated following adjustment for IGF-I. Using data from BCAC, the multivariable MR OR for overall BCa per SD increase in TG, conditioned on IGF-I, was 0.92 (95%CI 0.86, 1.10; *p* = 0.03). In UKBB, estimates for the effect of TG on overall BCa after conditioning on IGF-I were comparable. When results from BCAC and UKBB were meta-analysed, the multivariable MR OR for overall BCa per SD increase in TG, conditioned on IGF-I, was 0.93 (95%CI 0.88, 0.99; *p* = 0.03), which did not differ in size or magnitude when compared to the IGF-I unadjusted model (OR 0.94; 95%CI 0.89, 1.00; *p* = 0.05) (**Table 6**).

**Table 6.**
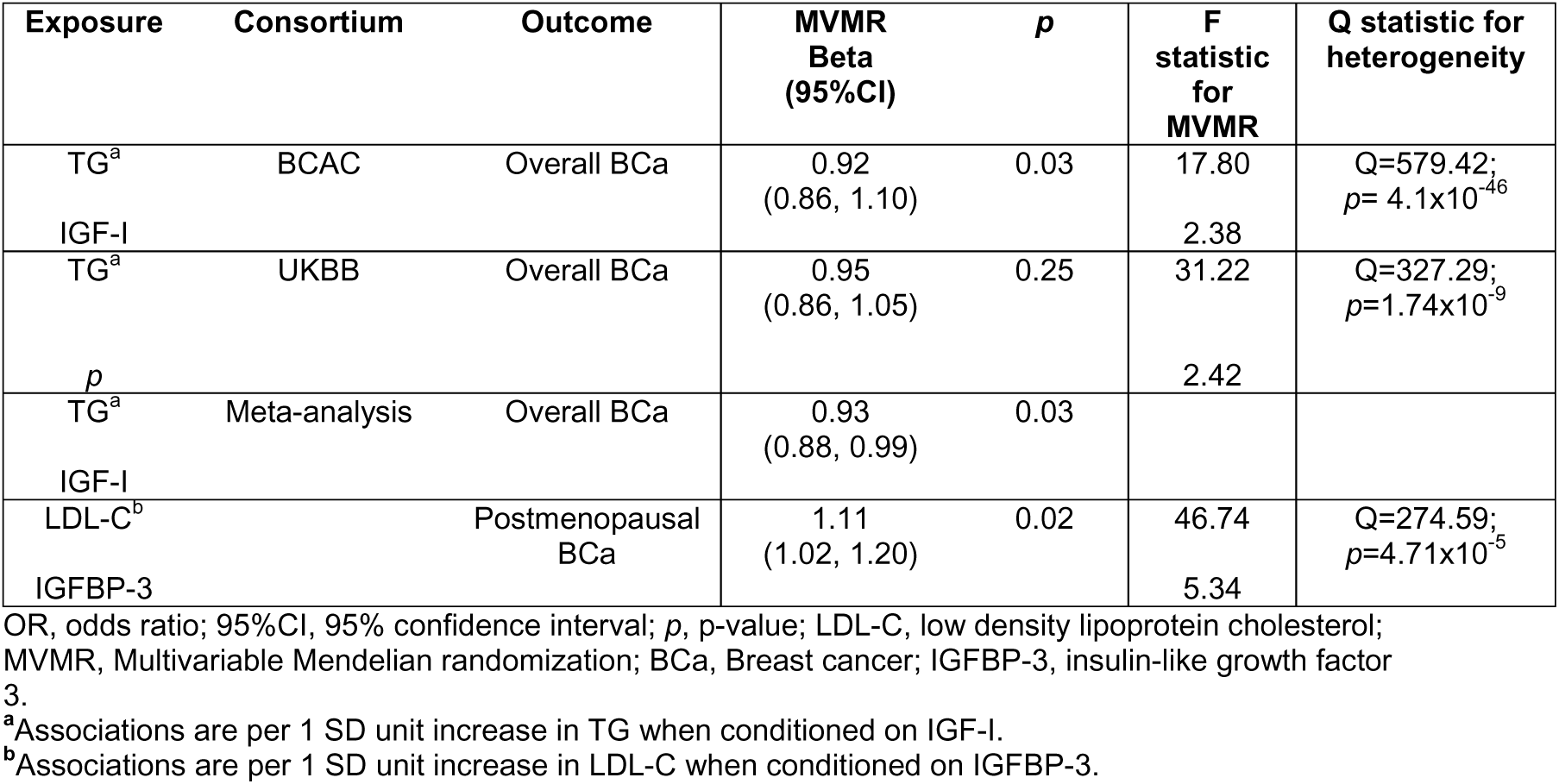
Multivariable MR analysis of the association of LDL-C and TG with breast cancer conditioned on IGFs.

Using data from UKBB, MVMR analyses were conducted to investigate if the effect of LDL-C on post-menopausal BCa was attenuated following adjustment for IGFBP-3. The multivariable OR for postmenopausal BCa per SD increase in LDL-C, conditioned on IGFBP-3 was 1.11 (95%CI 1.02, 1.20; *p* = 0.02), which did not differ by size and direction of effect when compared to the IGFBP-3 unadjusted model (OR 1.13; 95%CI 1.04, 1.25; *p* = 0.004) (**Table 6**).

### Estimating the causal relationship between LDL-C raising genetic variants in *HMGCR* with BCa and IGF levels

LDL-C raising variants in *HMGCR* (which simulate activation of *HMGCR*) were associated with increased odds of BCa in BCAC (OR 1.20, 95%CI 1.02, 1.40; *p* = 0.03). In UKBB, the estimated causal effect of LDL-C raising variants in *HMGCR* was consistent but less precise (OR 1.34, 95%CI 0.90, 2.02; *p* = 0.15). The meta-analysed OR for overall BCa in BCAC and UKBB was 1.22 (95%CI 1.04, 1.43; *p* = 0.01) (**Table 7**). Using data from BCAC, LDL-C raising variants in *HMGCR* were associated with increased odds of ER-positive BCa (OR 1.21, 95%CI 1.00, 1.47; *p* = 0.05) but not ER negative BCa (OR 1.20, 95%CI 0.90, 1.60; *p* = 0.23).

**Table 7.** Estimates of the causal relationship between LDL-C raising variants in *HMGCR* with breast cancer or IGF levels.

| Exposure | Consortium | Outcome | N | OR<sup>a</sup><br>(95% CI) | p |
| --- | --- | --- | --- | --- | --- |
| **LDL-C**<br>**raising**<br>**genetic**<br>**variants in**<br>***HMGCR*** | BCAC | Overall BCa | 228,951 | 1.20<br>(1.02, 1.40) | 0.03 |
|  | UKBB | Overall BCa | 230,954 | 1.34<br>(0.90, 2.02) | 0.15 |
|  | Meta-analysis | Overall BCa | 459,905 | 1.22<br>(1.04, 1.43) | 0.01 |
|  | BCAC | ER positive BCa | 69,501 | 1.21<br>(1.00, 1.47) | 0.05 |
|  | BCAC | ER negative BCa | 21,468 | 1.20<br>(0.90, 1.60) | 0.23 |
|  | UKBB | Premenopausal BCa | 59,124 | 1.47<br>(0.57, 3.75) | 0.42 |
|  | UKBB | Postmenopausal BCa | 112,245 | 1.63<br>(1.06, 2.49) | 0.03 |
|  | IGF working group of<br>CHARGE consortium | IGF-I levels | 30,884 | 0.20<br>(-0.02, 0.43) | 0.07 |
|  |  | IGFBP-3 levels | 18,995 | 0.03<br>(-0.24, 0.30) | 0.82 |

OR, odds ratio; 95% CI, 95% confidence interval; p, p-value; LDL-C, low density lipoprotein cholesterol; *HMGCR*, HMG-CoA reductase; BCa, breast cancer; BCAC, breast cancer association consortium; UKBB, UK Biobank; IGF-I, insulin-like growth factor 1; IGFBP-3, insulin-like growth factor binding protein 3.
<sup>a</sup>Associations are per 1 SD unit increase in LDL-C as proxied by genetic variation in *HMGCR*.

Using data from UKBB, LDL-C raising variants in *HMGCR* were associated with increased odds of postmenopausal BCa (OR 1.63, 95%CI 1.06, 2.49; *p* = 0.03), but not premenopausal BCa (OR 1.47, 95%CI 0.57, 3.75; *p* = 0.42) (**Table 7**). There was weak evidence that statins could reduce IGF-I (beta: 0.02, 95%CI –0.02, 0.43, *p* = 0.07). There was little evidence that genetic variants in *HMGCR* were associated with IGFBP-3 (**Table 7**).

## Discussion

In this study, we aimed to estimate the effect of circulating lipids and IGF traits on BCa and to investigate the role of circulating IGFs in the association between circulating lipids and BCa. We used complementary approaches to investigate circulating IGFs as causal intermediates between lipids and BCa. We found that TG was inversely associated with IGF-I and overall BCa and that IGF-I is positively associated with overall BCa. We also found that LDL-C was positively associated with IGFBP-3 and postmenopausal BCa and that IGFBP-3 is positively associated with postmenopausal BCa. Two-step MR results supported a hypothetical pathway between TG, IGF-I and overall BCa and also between LDL-C, IGFBP-3 and postmenopausal BCa.

A recent two-sample MR study assessed the relationship between circulating lipids and BCa in BCAC^11^. In an attempt to account for pleiotropic effects, the authors conducted sensitivity analyses by excluding pleiotropic variants. However, manual pruning of SNPs that are considered pleiotropic may result in an instrument that does not account for the underlying genetic architecture of the trait, resulting in findings that can be challenging to interpret^52^. Given that genetic variants included in the model are associated with multiple lipid fractions, the MR assumption that all genetic variants are valid instruments (i.e. not pleiotropic) is unlikely to hold. In this investigation, we have employed a multivariable MR approach, which allows genetic variants to have pleiotropic effects on other variables included in the model and jointly estimates the independent causal effect of each risk factor on the outcome. Our multivariable MR analysis found that TG is negatively associated with overall BCa, which was not observed in previous MR studies^11,14^. Care is needed in interpretation however as whilst these may reflect real differences, they could also be because of differences in analytical complexities. For example, MVMR model estimates the direct effect of each lipid conditional on the other lipid traits which differ from the total effect of the lipids as estimated by univariable MR analyses in the absence of pleiotropy.

Epidemiological evidence supports a relationship between lipids and IGFs^15–17^. In the context of this relationship, we set out to assess the relationship between circulating lipids and IGFs using a combination of observational and MR methods. Results from observational analyses and MR analyses highlight that TG decreases IGF-I levels and that TG and LDL-C increases IGFBP-3 levels. There was minimal evidence to suggest that circulating IGFs causally affect lipid levels, although this finding may be limited by the availability of robust genetic instruments for individual IGF traits^53^.

A meta-analysis of 17 studies and a case-control study showed that circulating IGF-I increases odds of ER-positive BCa in pre- and postmenopausal women but not odds of ER-negative BCa^22,54^. This finding is concordant with results of our study, suggesting that circulating IGF-I increases overall BCa, likely driven by ER-positive BCa. Our MR results suggest that circulating IGF-I is more influential in ER-positive BCa. However, preclinical evidence suggests that IGF signalling is mitogenic for both ER-positive and ER-negative BCa^55^. As we have used genetic variants that predict circulating levels of IGF-I, we cannot rule out important aspects of tissue-specific regulation which may contribute to BCa.

Menopause causes substantial disturbances to lipid metabolism^56^. Observational evidence suggests that the association between lipids or IGFs and BCa depends on menopausal status^5,22^. Using data from UKBB, our MR results suggest that circulating LDL-C is more influential in postmenopausal BCa compared to premenopausal BCa. In contrast to previous studies^57–60^, the association between HDL-C and BCa did not differ by menopausal status in our analysis. IGFBP-3 appeared to be more influential in post-menopausal BCa, consistent with findings from a meta-analysis of observational studies^22^. However, confidence intervals were overlapping for both LDL-C and IGFBP-3 estimates in premenopausal and postmenopausal BCa analyses and the difference could be due to low statistical power in subgroup analyses, in particular the small number of cases in the premenopausal analysis (n = 1,781).

Our two-step MR results suggested a pathway between TG, IGF-I and overall BCa and a pathway between LDL-C, IGFBP-3 and postmenopausal BCa. However, multivariable MR analyses, wherein either IGF-I or IGFBP-3 were included as a covariate in the model, did not attenuate the associations between TG and BCa or the associations between LDL-C and postmenopausal BCa. This is counter to our interpretation of results, however our main concern in this sensitivity analysis relates to instrument strength and the power of this analysis. Instrument strength in a MVMR model can be assessed using a conditional F statistic that summarises the relationship between genetic instruments and each exposure conditional on the other exposures included in the MVMR model^48^. We assessed the instrument strength in the multivariable MR models and found that the conditional F statistics for IGF-I and IGFBP-3 were low (**Table 6**). In addition, assessment of the Q-statistic for heterogeneity found evidence of potential pleiotropy in the MVMR model (**Table 6**), however, this could be due to presence of conditionally weak instruments for the IGF measures.

Given that our study found evidence that LDL-C increases both BCa and IGFBP-3 levels, it was appropriate to assess how statins (the most commonly prescribed LDL-lowering therapy) impact BCa and IGFBP-3 levels using LDL-C raising genetic variants in *HMGCR*. Increased LDL-C as proxied by genetic variants in *HMGCR* was associated with increased overall BCa, consistent with results from a previous MR study^13^. In our analysis the effect estimate for overall BCa using the LDL-C raising genetic variants in *HMGCR* was larger than the effect estimate using the 185 SNP genome-wide LDL-C score. It seems likely that the difference in the results could reflect properties of the instruments employed. The larger number of genetic variants used to proxy LDL-C levels could capture pathways beyond *HMGCR*, therefore pleiotropy could influence the effect estimate. As the 5 SNPs in the *HMGCR* used in the statin analysis are weakly correlated, the multiplicity of associations due to LD between SNPs could bias the MR results^61^. However, we attempted to account for the LD between the SNPs in the *HMGCR* instrument in our MR analyses by accounting for these^49,50^. Differences in the results using the *HMGCR* score and 185 SNP genome-wide LDL-C score could also be due to off-target effects of statins (i.e. not via LDL-C). For example, a recent study by Kar et al^13^ suggested that BMI, rather than LDL-C, could be the true underlying mechanism responsible for the protective association between genetic inhibition of *HMGCR* and BCa.

Randomised control trials have shown that statin use results in altered levels of IGF-I and IGFBP-3^15–17^. Our study found weak evidence of an association between genetic variants in *HMGCR* and IGF-I levels and minimal evidence of an association with IGFBP-3 levels. Failure to detect an association between statins an IGFBP3 may be due to the relatively small size of the IGF GWAS.

## Conclusion

This study employed complementary observational and MR analyses to confirm that LDL-C and HDL-C are positively associated with overall BCa and that TG is negatively associated with overall BCa. Our study found that TG is negatively associated with IGF-I and that IGF-I is positively associated with overall BCa, consistent with the TG-BCa association. We also found that LDL-C is positively associated with IGFBP-3 and that IGFBP-3 is positively associated with postmenopausal BCa, consistent with the LDL-C-postmenopausal BCa association. Our two-step MR results build on evidence linking circulating lipids and IGFs with BCa, however, multivariable MR analyses are currently unable to support this relationship due to presence of conditionally weak instruments for the IGF measures.

## Data Availability

Data available on request from the authors

## Acknowledgements

Quality Control filtering of the UK Biobank data was conducted by R.Mitchell, G.Hemani, T.Dudding, L.Paternoster as described in the published protocol (doi:10.5523/bris.3074krb6t2frj29yh2b03×3wxj). SHIP is part of the Community Medicine Research net of the University of Greifswald, Germany, which is funded by the Federal Ministry of Education and Research (grants no. 01ZZ9603, 01ZZ0103, and 01ZZ0403), the Ministry of Cultural Affairs as well as the Social Ministry of the Federal State of Mecklenburg-West Pomerania, and the network ‘Greifswald Approach to Individualized Medicine (GANI_MED)’ funded by the Federal Ministry of Education and Research (grant 03IS2061A).

## Tables and Figures

Supplementary Table 1. Genetic instruments used to construct genetic risk scores for lipid traits used in multivariable MR analyses

Supplementary Table 2. Genetic instruments used to construct genetic risk scores for IGF-I and IGFBP-3

Supplementary Table 3. Study characteristics of the SHIP cohort

Supplementary Table 4. Association of IGF axis and lipids with potential confounders in the SHIP study

